# Travel distance to the general practitioner. Do patients move closer to the services when starting to use them?^1^

**DOI:** 10.1101/2022.08.31.22279204

**Authors:** Rosanna N. I. Johed, Kjetil Telle

## Abstract

**Objectives:** The main objectives were 1) to calculate and describe travel time by car from the home of Norwegian residents to the office of their named general practitioner (GP); 2) to estimate changes in travel time for residents who started to visit the GP and, if so, to 3) to explore if the residents changed GP or moved to reduce their travel time.

**Methods:** We used nation-wide individual-level annual registry data 2009-2017 on the exact location of the home of every resident and the GP-office to calculate travel time in minutes by car from home to their assigned GP. First, using data for 2017 only, we calculated travel time at the median and 90^th^ percentile, and by sex, age, immigrant background, county of residence and use of GP in 2017. Second, with annual data 2009-2017, and restricting the sample to residents who had not used their GP over the last two years (*t-2* and *t-1*), we used a difference-indifferences model to estimate changes in travel time in the next two years (*t+1* and *t+1*) for patients with a visit in year *t* compared with those with no visit in *t*. Separate models were run for those who changed GP and those who moved from *t-2* to *t*., and for the 20% who lived farthest away in *t-1*.

**Results:** We could calculate the travel time for 3,976,910 residents in 2017, with a median travel time from home to the GP of 4.9 minutes and a travel time at the 90^th^ percentile of 18.3 minutes. In the most sparsely populated northern county of Norway, travel time was about 5 minutes at the median and below 45 minutes at the 90^th^ percentile. Elderly residents and residents who used their GP in 2017 had a somewhat shorter travel time than other groups of the population. Using annual data for 2009-2017 in the difference-in-differences analysis (16,388,151 resident-year observations), travel time dropped by 2.5 minutes (95% confidence interval 2.4 to 2.6) in *t+1* and *t+2* for patients with a visit in *t* compared with similar patients with no visit in *t*. The drop was similar for patients who did and did not change GP, but larger for patients who moved (10.0 minutes; 95%CI 9.7 to 10.4) compared with those who did not move (0.6 minutes; 95%CI 0.5 to 0.7), and particularly large for the 20% living farthest away in *t-1* (24.2 minutes; 95%CI 23.3 to 25.2).

**Conclusions:** Travel time from home to ones GP is short for the vast majority of the population in the sparsely populated country of Norway. However, residents move closer to the GP when they start using the services, especially patients who used to live far away. This relocation may reflect strong preferences for proximity to the services, and we conclude that more knowledge is needed to enable transparent balancing of costs and benefits of centralizing GP-services, at least in rural areas.

**JEL classification:** I10, E32, J6

## Introduction

It is well-documented that sociodemographic factors affect access to health care services of high-quality (Fiva et al. 2014, Van Doorslaer et al. 2000, Vikum et al. 2012), and also that longer travel distance to the services is associated with both lower utilization and worse health (Celaya et al. 2012, Kelly et al. 2016, Ludwick et al. 2009, Nemet and Bailey 2000, O’Reilly et al. 2001, Raknes et al. 2012). However, little is known not only about causal relationships between travel distance and utilization, but even about how far from the services residents actually live. A few earlier studies have had access to precise measures of travel time or distance from the full home address of the included patients and the health services, but most have used self-reported travel time or aggregated measures of start and end location (Kelly et al. 2016). For example, Raknes et al. (2013) used distance from the municipality population centroid of the patients to the nearest casualty clinic for ten municipalities, with the obvious drawback that some patients can live far from the centroid. None of the few longitudinal studies included in the review of Kelly et al. (2016) accounted for changes in travel distance from residential relocation as people moved house over time. While there are many survey-based studies of health-related residential relocation in various groups of patients (Wilmoth 2010), we are not aware of any nation-wide longitudinal study of residential relocation for all inhabitants who start to use general practitioner (GP) services after a few years of non-use.

Thought the more interesting question is how travel distance affects utilization, and ultimately treatment quality and health, it is also important to provide more knowledge about actual distance to the services for various groups of the population and how it may affect utilization and residential relocation when starting to use the services. Indeed, the geographical location is often central in debates about the organization and centralization of health care services (see e.g. Fischer et al. 2022), and where to locate services is largely under the discretion of public policy regulation and decision. If patients who start to use the services after a time of non-use are taking on the costs of moving closer to them, this may reflect how proximity is valued by the patients, and is thus relevant knowledge for policy regulation and decision.

In contributing knowledge to such debates and policies, our aim was to calculate and describe travel time by car from the home of all Norwegian residents to the office of their assigned GP, and to see if the residents changed GP or moved to reduce their travel time when starting to visit the GP.

## Methods

### Study setting

To reap the benefits of continuity of care, every resident of Norway is listed with one named regular general practitioner (GP) who is responsible for taking care of their medical needs, including referrals to specialist care (Sandvik et al. 2022). Thus, the GP is central for access to most of the high-quality, universally accessible and publicly funded health services in Norway. Residents can change GP up to twice a year. Almost all health services (except dental) are funded by the governmental health insurance, and out of pocket expenses are either non-existent or very low by Norwegian standards (less than 20-30US$ for a consultation to the GP, and all expenses for health services or prescribed drugs exceeding a ceiling of less than 300 US$ a year are covered fully by the insurance). Membership of the insurance is compulsory for all residents, costs are covered by general taxes, and the quality and funding of the publicly-funded (but often privately-provided) health services are so generous that privately funded alternatives are very rare. In 2018 there were just below 4,800 GPs in Norway, covering a population of 5.3 million, with more GPs per resident in rural than urban areas (SSB 2019).

With about 14 residents per square kilometer in 2017, Norway is parsley populated compared with European countries like Germany (237), France (122) and Denmark (144), but more similar to the United States (36), Sweden (25), Finland (18) and Canada (4) (WorldBank 2022). However, there are large differences in population density across Norway’s 19 counties at the time, with about 1500 residents per square kilometer in the capital of Oslo and below 2 in the northernmost and most sparsely populated county of Finnmark (SNL 2022).

### Data sources

The Population Registry maintained by Statistics Norway including individual level information on demographics of all Norwegian residents was combined with the General-Practitioner Registry (maintained by the Norwegian Directorate of Health) containing the GP assigned to each resident, to obtain individual-level annual data for the years 2009-2017. The location of the home of every resident was also available in the population registry and the location of the GP-offices was retrieved from the Norwegian Registry of Business Enterprises. Age, sex, immigration background and county of residence was retrieved from the population registry, and consultations with the GP was available from the National Reimbursement Register that covers consultations between the GP and the resident.

Information was linked across the registries and over time using the unique personal identification number (a project-specific encrypted version) provided every resident of Norway at birth or upon immigration (similarly for the unique enterprises identifier of the GPs). In general, the quality and completeness of these registries are considered to be very good (Bakken et al. 2020, Lyngstad and Skardhamar 2011, Røed and Raaum 2003).

### Travel time and other variables

Using the coordinates of the home address of each individual and of office of the GP, we calculated the minimum travel time in minutes by car between the two locations. In practice, we divide Norway into 100-meters squares collecting all residents living within each square and likewise for the GP-offices. The shortest travel time in minutes from the center of the home-square to the center of the GP-office-square by car was calculated using the most updated map of roads (with speed limits) at the given year, as provided by the Norwegian Mapping Authority, using the software ArcGIS Pro for the calculations. For a few distances the algorithm was obviously astray, and to handle extreme outliers we set travel times above the 99^th^ percentile to the value at the 99^th^ percentile.

Sex was included as a binary variable, and so was also immigrant background (including immigrants and their Norwegian-born children; and the rest of the population). Age was operationalized in six categories (0-19, 20-29, 30-49, 50-66, 67-79, 80 and above), and county of residence in 19 categories (one for each of Norway’s 19 counties at the time). Visit to the GP includes face-to-face consultations, and operationalized in three categories: no visit, one visit, or more than one visit during the calendar year.

### Study population

For the cross-sectional analysis we started out with data on every resident of Norway in 2017. However, a substantial number of GP-offices could not be located with coordinates, and the residents of these GPs were thus excluded from the analysis (see Appendix for robustness check). We also dropped a small proportion of residents who did not have a GP at the time or for whom we were unable to calculate the travel time to their GP (see Appendix for details). The sample for the cross-sectional analysis in 2017 comprised 3,976,910 residents.

For the longitudinal analysis we combined datasets like the one for 2017 for all available years 2009-2017. To operationalize a starting point for using of GP-services, we kept only the residents who had not visited the GP for two consecutive years *t-2* and *t-1* (e.g. 2009 and 2010), and split them in two groups according to whether they visited (“treated group”) the GP or not (“comparison group”) in year *t* (e.g. 2011). The travel time in minutes to the GP was traced over the five years from *t-2* through *t+2* (e.g. 2009, 2010, .., 2013) for each resident. I.e. we traced the travel time of residents over 2009-2017 and compared its development across residents i) who started to use the GP after two years of non-use and ii) who did not start to use the GP in the third year. The sample for the longitudinal analysis 2009-2017 comprised 16,388,151 resident-year observations.

### Statistical analyses

First, we explored travel time in 2017 descriptively by groups of age, sex, county of residency, immigrant background and same-year utilization of GP services. For each group, results were illustrated in graphs with percentiles of travel time on the x-axis (for every 10^th^ percentile) and travel time in minutes on the y-axis.

Second, we studied the change in travel time for those who had not visited the GP for at least two years (*t-2* and *t-1*) and then 1) visited the GP in year *t* and 2) still did not visit the GP in year *t*. In estimating changes in travel time across these two groups, we used a difference-in-differences (DiD) approach. DiD analyses evaluate the effect of an event by comparing the change in the outcome for the affected group before and after the event, to the change over the same time span in a group not affected by the event (Dimick and Ryan 2014, Angrist and Pischke 2009, Wing et al 2018). In this study, we compared the travel distance in the two years before and after the resident did (difference 1) or did not (difference 2) start to visit the GP. The DiD estimate is the difference between these two differences, estimated using linear probability models with robust standard errors (clustered at the individual level) and presented as a difference in minutes. By including calendar year fixed effects, this approach accounts for background trends, like increased use of electronic consultations, new regulations, procedures or drugs, or improved road quality. The DiD estimate can be interpreted as the change in travel time that is related to the starting of visiting the GP, beyond any shared temporal trends. If there is no relationship between starting to visit the GP and subsequent travel time, the DiD estimate would be zero.

We adjusted for the following individual characteristics: Sex (boy/girl), age groups (0-19, 20-29, 30-49, 50-66, 67-79, 80 and above), county of residence (19 categories), immigrant background (yes/no) and calendar year (2009, 2010, .., 2017). Models were run separately for residents who did and did not switch GP (operationalized as having a GP in another 100-meters square) or move (operationalized as living in another 100-meters square) from t-2 to t, and for the 20% with the longest travel time in t-1 (vs. the rest). For robustness we also looked at some analyses when we imputed missing GP-locations and when we used Pythagorean straight lines (in kilometers) instead of travel time (see Appendix). All analyses were run in STATA MP v.16. The project was approved by The Regional Ethics Committee in Norway (#2017/373).

## Results

In the dataset of all 3,976,910 residents of Norway in 2017 for whom we could calculate the distance to their GP (see Appendix), 50% were women and the mean age was 40.1 years (Appendix Table A1). 16.8% had immigrant background. 11% of the residents lived in the most densely populated county of Oslo, and 1% in Norway’s most sparsely populated and northernmost county of Finnmark. 34 % did not visit the GP in 2017, 19% visited once and 47% more than once.

The median travel time from home to the GP was 4.9 minutes, and the travel time at the 90^th^ percentile was 18.3 minutes; see Figure 1. Men had a somewhat longer travel time (median 5.0; 90^th^ percentile 19.1) than women (median 4.8; 90^th^ percentile 17.5); Figure 2.

**Figure 1:**
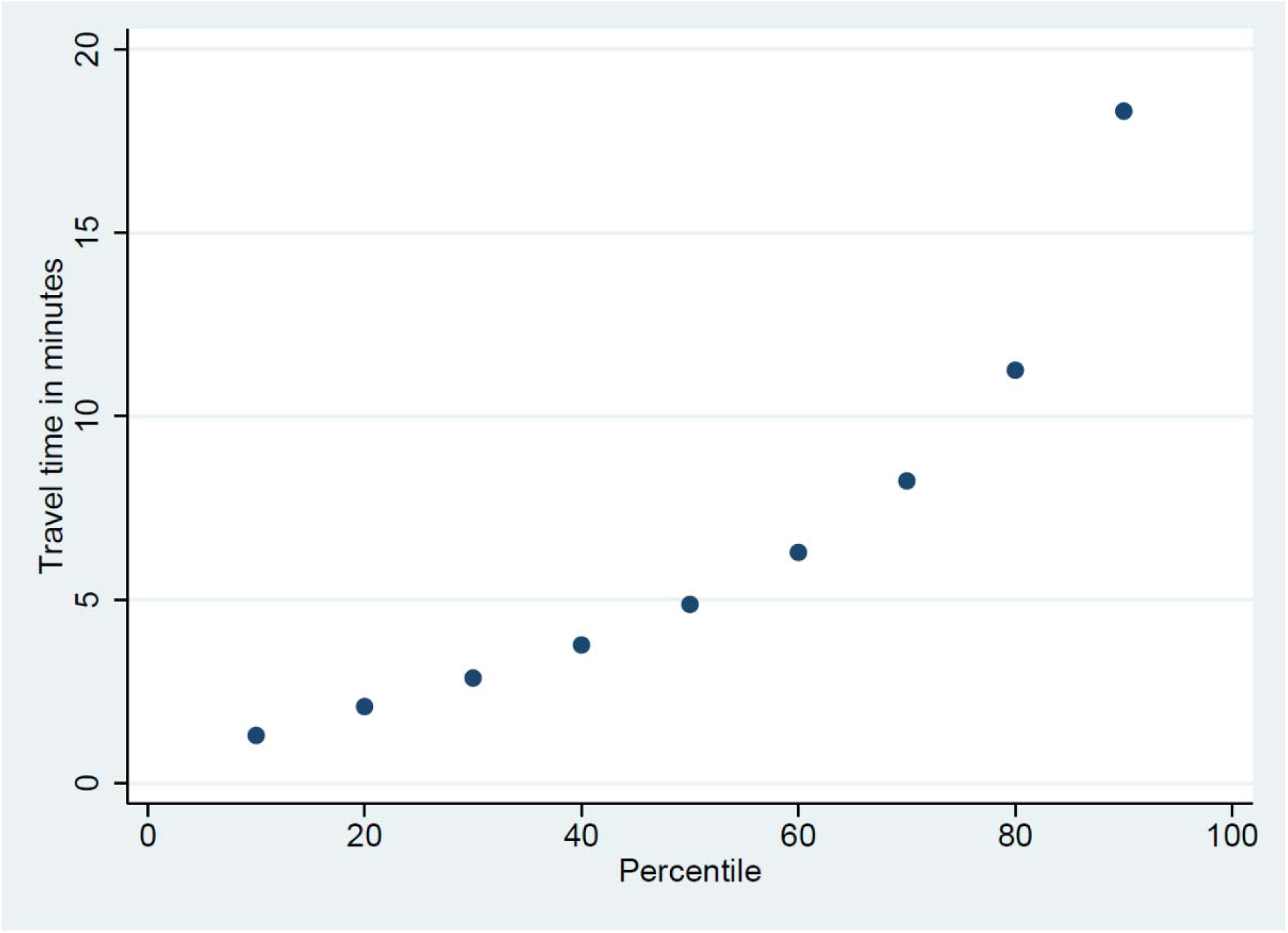
Travel time in minutes from home to the general practitioner by car for the 1^st^ to the 9^th^ decile of Norwegian residents. 2017.

**Figure 2:**
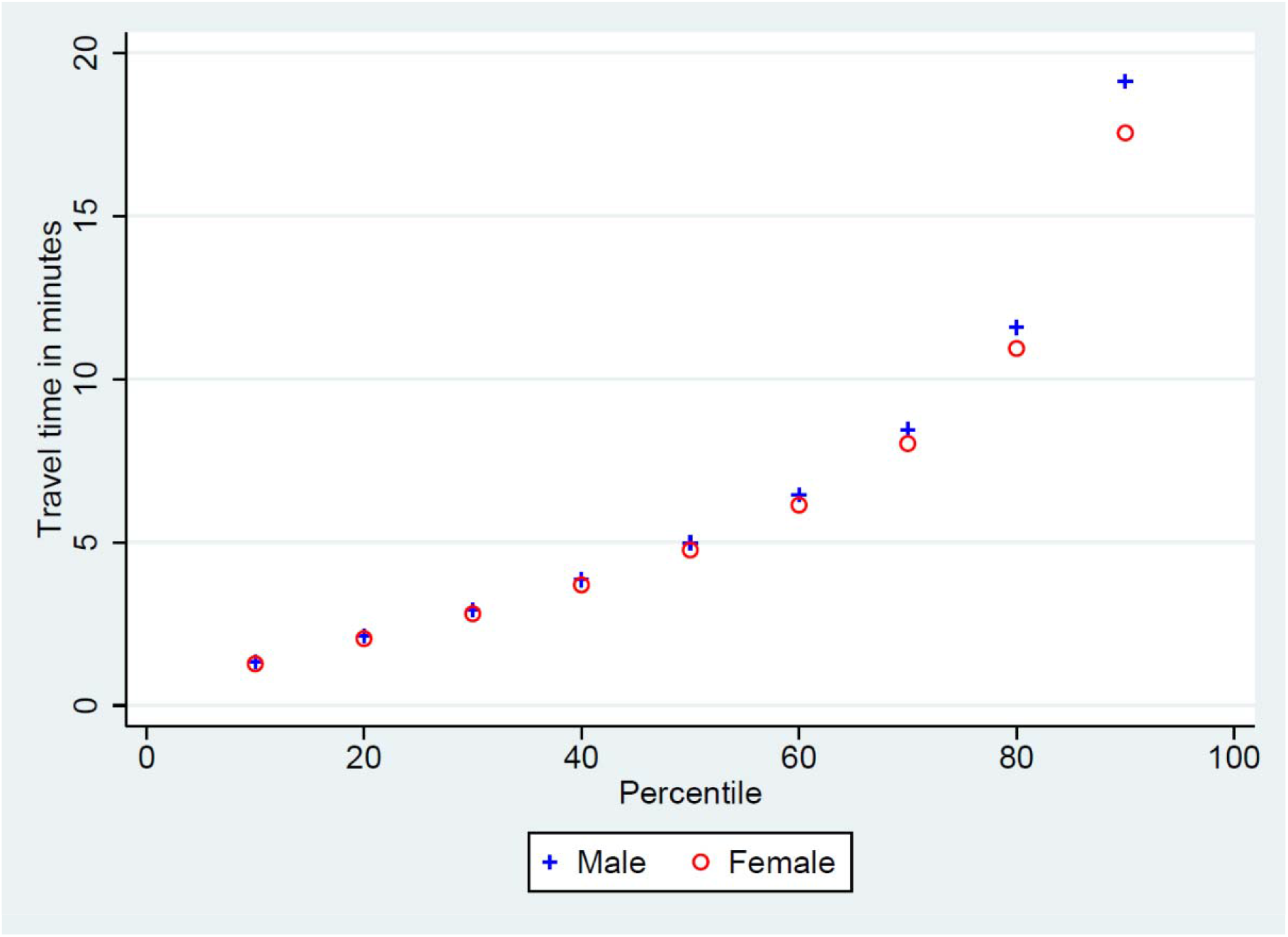
Sex differences in travel time in minutes from home to the general practitioner by car for the 1^st^ to the 9^th^ decile of each group. 2017.

In general travel time tended to be lower the older the resident for age groups above 30 years (Figure 3). Calculated travel time for young adults aged 20-30 years was much higher in the upper part of the distribution of travel time (median 5.5; 90^th^ percentile 76.0) than for the rest of the population (Figure 3).

**Figure 3:**
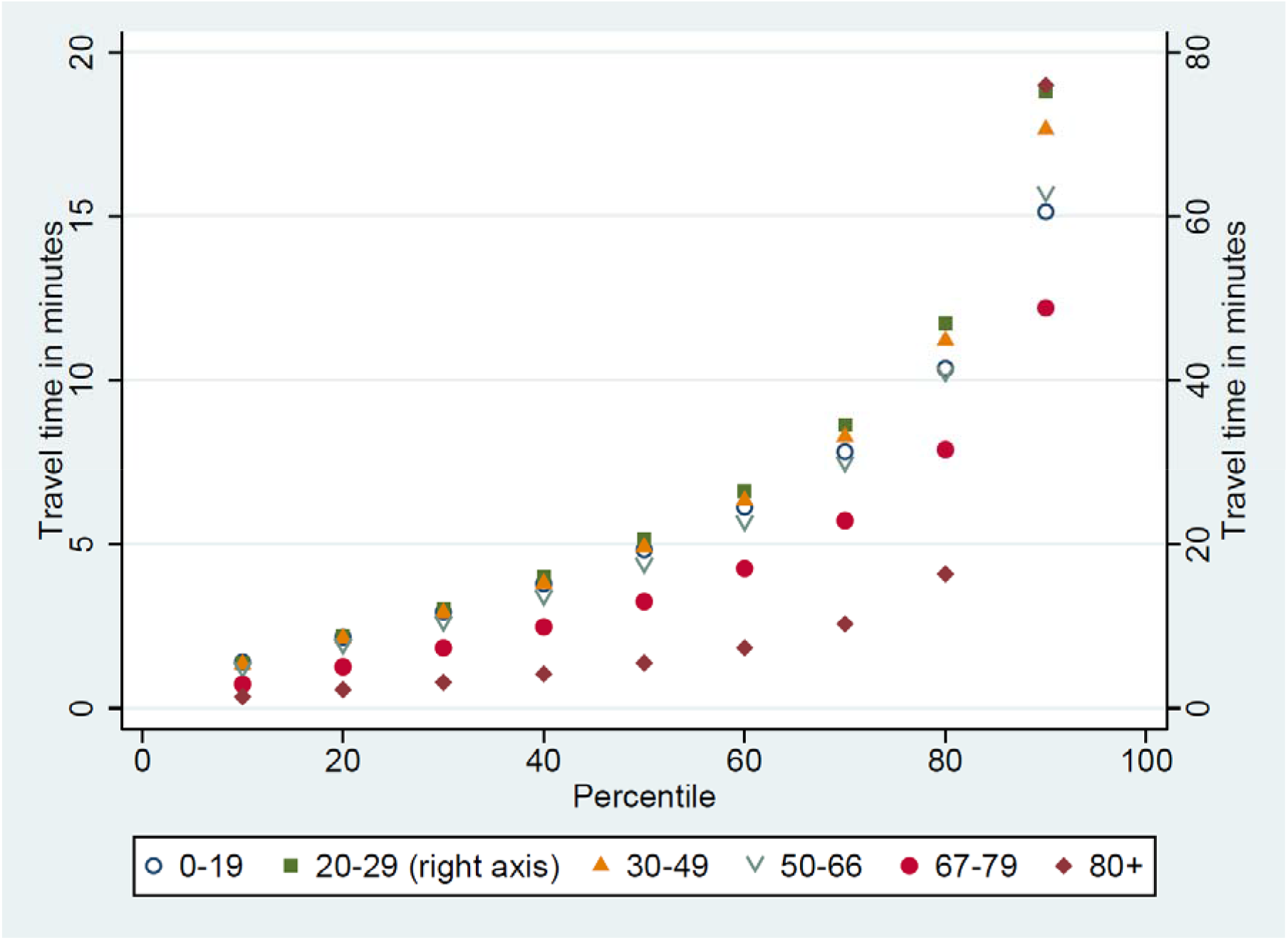
Age differences in travel time in minutes from home to the general practitioner by car for the 1^st^ to the 9^th^ decile of each group. 2017.

While the median travel time was similar across all of Norway’s 19 counties (Figure 4), the travel time at the 90^th^ percentile was higher in the most sparsely populated county in the north of Norway (44.1 minutes) than in the densely populated capital of Oslo (12.5 minutes). Residents with an immigrant background (median 4.3; 90^th^ percentile 16.9) had somewhat shorter travel time from home to the GP than the rest of the population (Figures 5).

**Figure 4:**
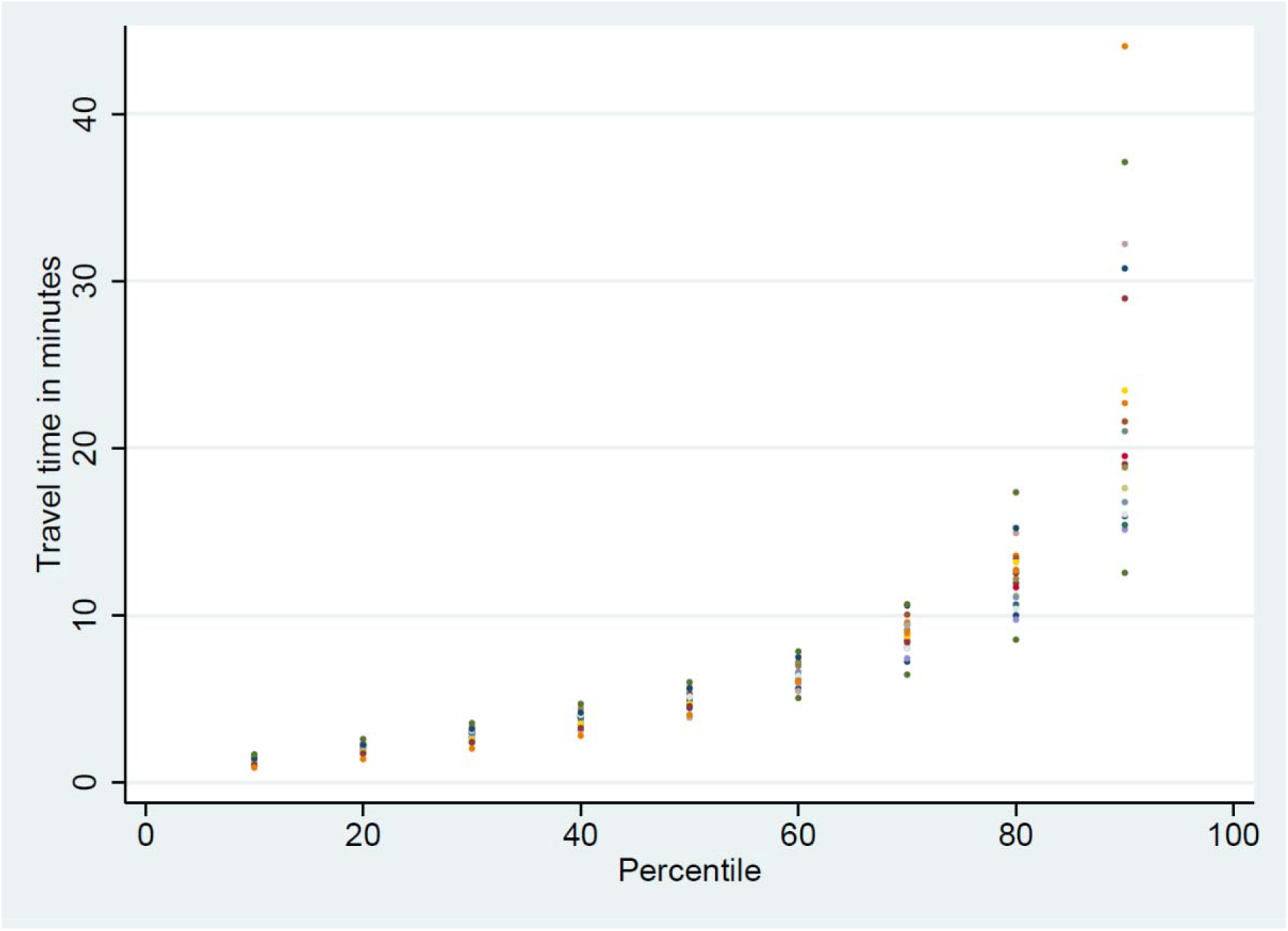
Differences by county of residency, in travel time in minutes from home to the general practitioner by car for the 1^st^ to the 9^th^ decile of each group. 2017. Note: One dot for the distribution of travel time in each of the 19 counties of Norway.

**Figure 5:**
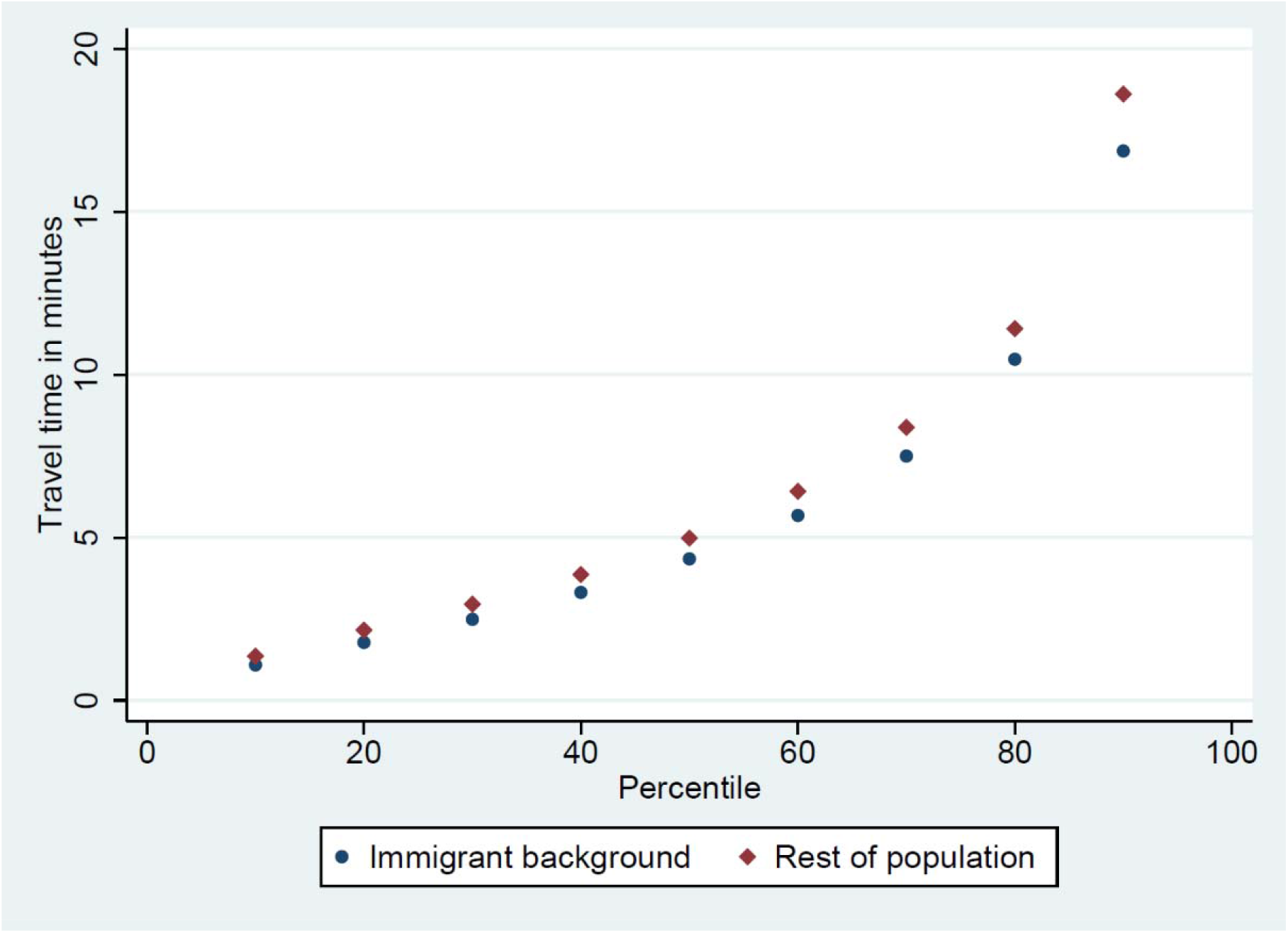
Differences by immigrant background, in travel time in minutes from home to the general practitioner by car for the 1^st^ to the 9^th^ decile of each group. 2017.

The residents who did not visit the GP in 2017 had longer travel time (median 5.0; 90^th^ percentile 21.5) than those who had one (median 4.9; 90^th^ percentile 17.7) or more than one (median 4.8; 90^th^ percentile 16.8) visit in 2017 (Figure 6).

**Figure 6:**
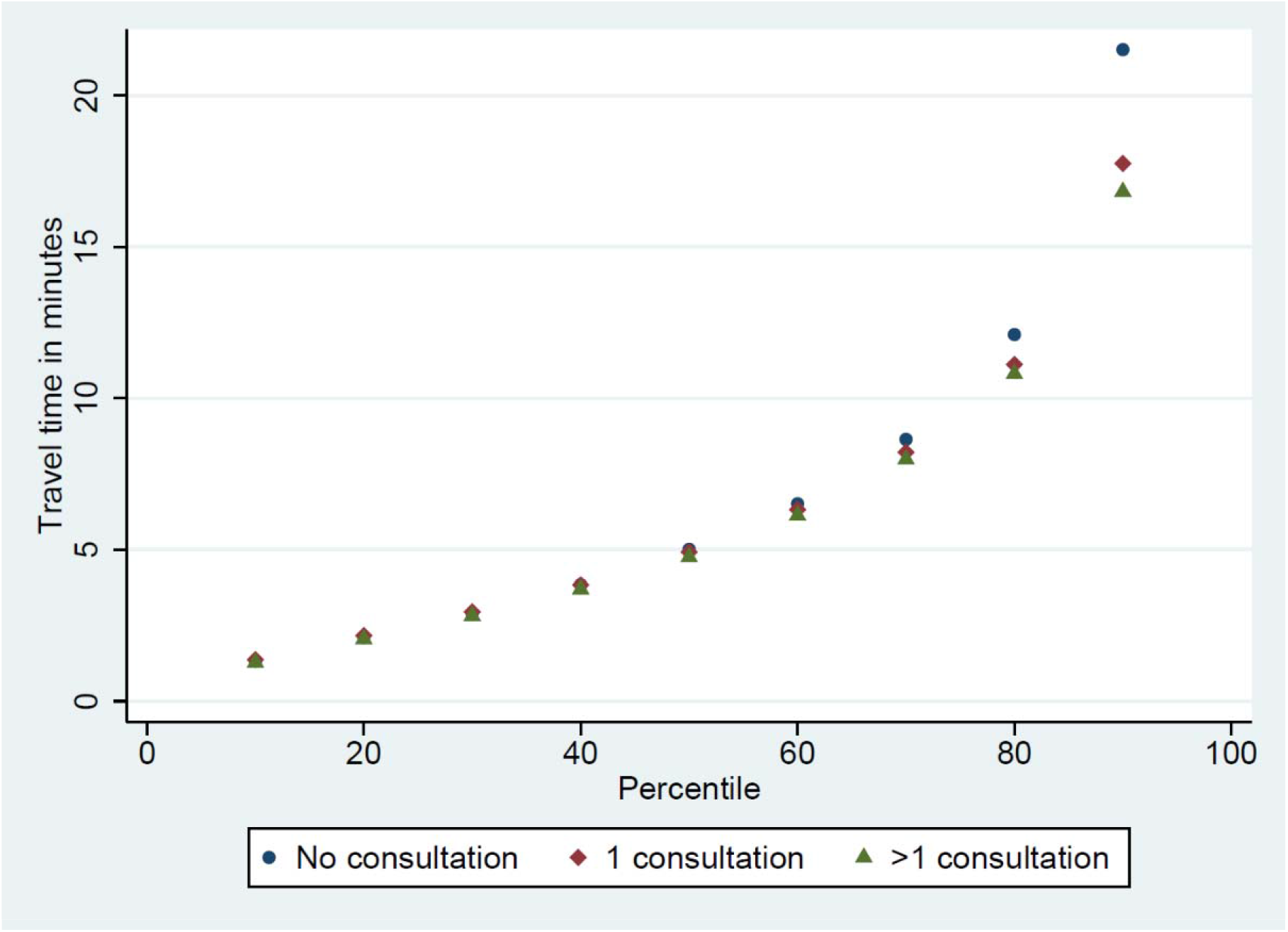
Differences by use of the general practitioner, in travel time in minutes from home to the general practitioner by car for the 1^st^ to the 9^th^ decile of each group. 2017.

The dataset following residents over 2009-2017 who did not visit the GP for two years (*t-2* and *t-1*), and then did or did not visit the GP in the third year (*t*), contained 16,388,151 resident-year observations. For those visiting the GP in *t*, travel distance dropped from 17.4 minutes in *t-1* to 14.4 minutes in *t+1*, while it remained similar (19.8 to 20.1) for those who still did not visit the GP in *t* (Figure 7). The difference-in-differences regression analysis thus shows that the travel time fell by 2.5 minutes (95% confidence interval 2.4 to 2.6 minutes) over *t+1* and *t+2* for those who started to visit the GP in *t* compared with those who did not (Table 1, row 1). The fall in travel time was 11.8 minutes (11.4 to 12.0) for the 20% of the residents who lived farthest away from the GP in *t-1* compared with 0.8 minutes (0.7 to 0.9) for the rest of the residents (Table 1, rows 2 and 3).

**Figure 7:**
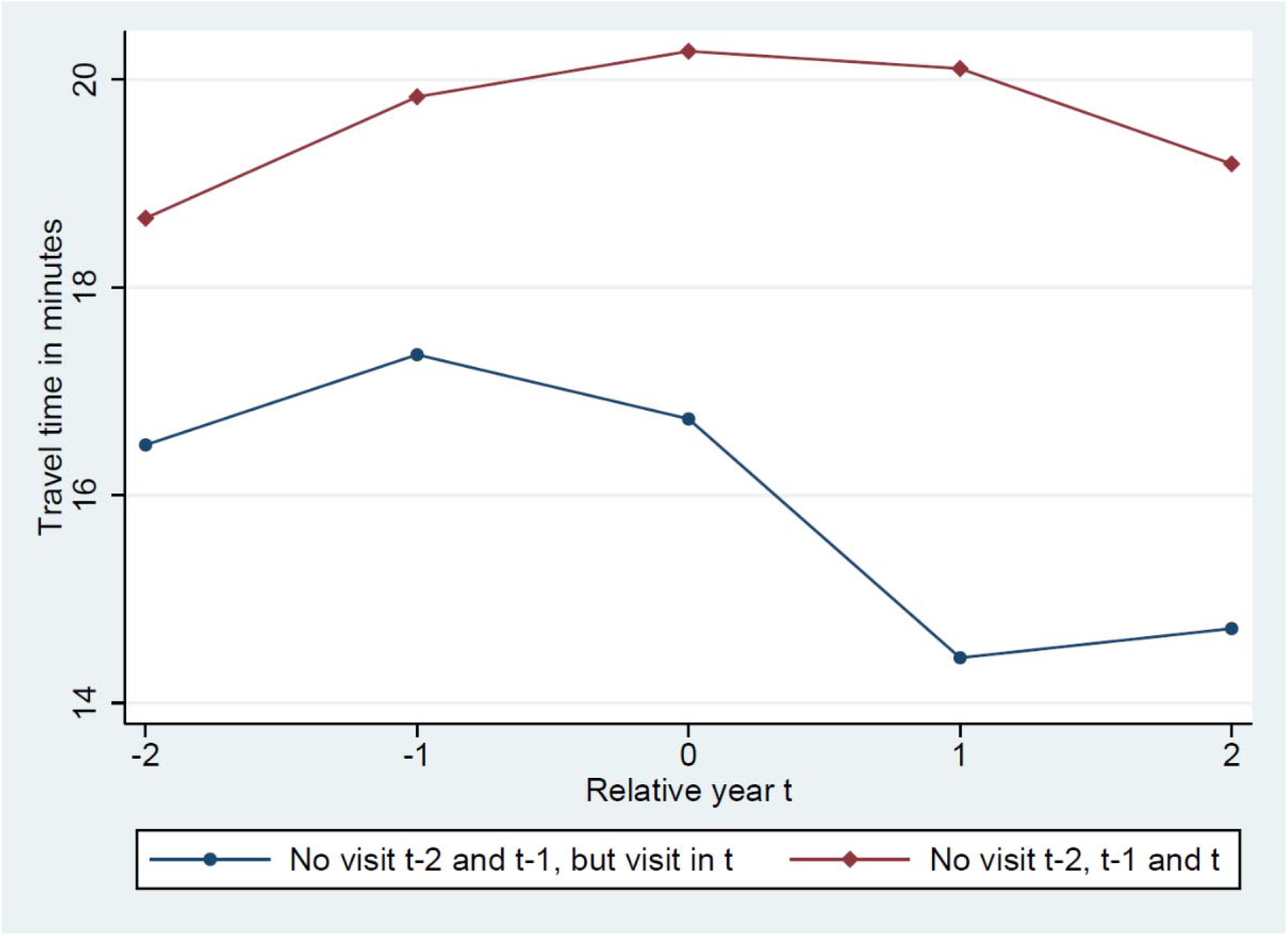
Mean travel time in minutes from home to the general practitioner by car for residents who had not visited the general practitioner for two years (*t-1* and *t-2*), and then i) visited the general practitioner in year *t*, or ii) did not visit the general practitioner in year *t*. Data for calendar years 2009-2017.

**Table 1:** Difference-in-differences estimates of the change in travel time to the GP in minutes for those who did visit the GP in *t* relative to those who did not visit the GP in *t*. The sample contains only residents who did not visit the GP in years *t-2* and *t-1*, and the longitudinal data were available for each year 2009-2017.

|  | DiD-estimate | 95% CI | N |
| --- | --- | --- | --- |
| Full sample | -2.5 | -2.6, -2.4 | 16,388,151 |
| *Long travel distance in t-1 (top 20%)* | -11.8 | -12.0, -11.4 | 3,252,311 |
| *Not long travel distance in t-1 (not top 20%)* | -0.8 | -0.9, -0.7 | 13,135,840 |
| Changed GP from t-2 to t | -2.1 | -2.7, -1.5 | 1,094,919 |
| Did not changed GP from t-2 to t | -2.1 | -2.2, -2.0 | 15,293,232 |
| *Moved from t-2 to t* | -10.0 | -10.4, -9.7 | 3,082,762 |
| *Did not move from t-2 to t* | -0.6 | -0.7, -0.5 | 13,305,389 |
| Moved from t-2 to t and had long travel distance in t-1 (top 20%) | -24.2 | -25.2, -23.3 | 890,834 |
| Moved from t-2 to t and did not have long travel distance in t-1 (not top 20%) | -5.6 | -5.9, -5.3 | 2,191,928 |

Note: Each row contains results from one separate regression model. In addition to the reported difference-in-differences estimate, all regression models included a variable with two categories for the group who started to use GP in $t$ , for the post period ( $t+1$ and $t+2$ ), for sex and for immigrant background, as well as a variable with several categories (see Methods Section for details) for calendar year, for age group and for county of residence. Robust standard errors clustered on the individual level were used to calculate the 95% confidence intervals (95% CI).

The fall in travel time for the residents who changed GP from *t-2* to *t* was 2.1 minutes (1.5 to 2.7), and it was also 2.1 minutes (2.0 to 2.2) for the residents who did not change GP (Table 1, rows 4 and 5). The fall in travel time for the residents who moved from *t-2* to *t* was 10.0 minutes (9.7 to 10.4) and it was 0.6 minutes (0.5 to 0.7) for the residents who did not move (Table 1, rows 6 and 7). Among those who moved, the fall was 24.2 minutes (23.3 to 25.2) for the 20% who lived farthest away from the GP in *t-1* compared with 5.6 minutes (5.3 to 5.9) for the rest of the residents who moved (Table 1, rows 8 and 9).

## Discussion

### Principal findings

In the sparsely populated country of Norway, the travel time from home to the general practitioner (GP) was less than 5 minutes at the median and less than 20 minutes at the 90^th^ percentile in 2017. Even in the most sparsely populated county of Norway, with less than two residents per square kilometer, median travel time was about 5 minutes and travel time at the 90^th^ percentile was below 45 minutes. Elderly residents had somewhat shorter travel time than other adults above the age of 30, and residents who used the GP in 2017 also had a shorter travel time than residents who did not.

When tracing the travel time of residents over 2009-2017, we found that it fell by 2.5 minutes for residents who started to use the GP after two years of non-use, compared with those who did not start to use the GP in the third year. The fall was driven by residents who moved closer to the GP (fell by 10 minutes), and in particular by the 20% of them who used to live farthest away from the GP (fell by 24 minutes).

### Interpretation and comparison to related studies

It is a commonly found empirical pattern that people who live farther away from the health services have decayed health compared with those who live closer by (Kelly et al. 2016). This “distance decay association” have spurred concerns that long travel distance can be a barrier to necessary utilization of health services, and some previous studies have found that people living farther away from the services use them less than those living closer by (Nemet and Bailey 2000, Ludwick et al. 2009, O’Reilly et al. 2001, Raknes et al. 2013). For methodological reasons, reliable evidence on causal relationships between travel distance and utilization are scarce (see though, e.g. Fischer et al. 2022 on closure of birth clinics), but even descriptive statistics on travel time for larger groups of residents to health services, and definitely at the country-level, are very rare (Kelly et al. 2016). Such knowledge of time to services can be important in policy decisions on organization and centralization of health services. For example, if travel time is very short for almost all residents, financial or organizational changes that is expected to result in further centralization may be of limited concern, while further centralization may not be politically acceptable if it affects parts of the population with long travel distance and particular needs.

We find that more than half of the Norwegian population have a travel time to the GP of less than 5 minutes, which appears short in the sparsely populated country. It is, however, in may not be that different from the findings by Raknes et al. (2013), who looked at the far more centralized emergency room services (Legevaktutvalget 2021) in 10 municipalities in the southern part of Norway. Though they used the travel time from the population centroid of the municipalities to the emergency room service, which might understate actual travel time for some inhabitants, they report a mean travel time of less than 6 minutes.

The population density across Norway’s 19 counties differs substantially, with about 1500 inhabitants per square kilometer in the capital of Oslo and below 2 in the northernmost and most sparsely populated county of Finnmark (SNL 2022). Still, median travel is similar across the Norwegian counties (5 minutes), though the travel time in the upper tail (90^th^ percentile) is higher in the most sparsely populated counties. The relatively small differences in travel time across Norwegian counties is reflecting that the number of GPs per resident are higher in rural than urban areas (SSB 2019). Thus, it is not completely clear that the findings for the sparsely populated country of Norway differ substantially from what one would find in other OECD-countries, since they also depend on residential patterns and how the primary health care services are organized.

With less than 5 minutes from home to the GP at the median, it seems unlikely that some further centralization of the services will have a substantial impact on utilization for this group. Thus, increasing travel distance may not be a serious concern with a reform that, say, contributed to more GPs sharing premises. Among population groups with long travel distance to the services, however, impacts of such reforms may be expected to increase costs of utilization too much.

As noted, the distance decay association may of course not reflect causal relation, i.e. that an increase in distance will reduce utilization. We find that those who start visiting the GP are moving closer to the GP, possibly reflecting a causal direction going, at least partly, from need of services to residential re-location closer to the GP. We note that such a temporal pattern of migration to health services when the services are needed, would show up in cross-sectional data (including in our Figure 4 for the 2017) as a distance decay association, as those using the services more have moved closer to them.

Health-related residential relocation is a well-known phenomenon (Wilmoth 2010, van der Pers et al. 2018). Of course, shorter travel time to the GP may coincide with access to other health-related amenities, like specialist services, pharmacies, easily maintained condominiums instead of an older house in the countryside, closeness to neighbors and relatives, transportation services, etc. Still, the substantial relocation effect we estimate, especially for the patients who used to live far away from their GP, may reflect strong preferences for proximity to such amenities when starting to use health services, making more knowledge of these preferences important for transparent balancing of costs and benefits of centralizing GP-services in rural areas. More knowledge of motivations for residential relocation will be important for making policy-decisions about the degree of centralization of health services, especially given the growing proportion of elderly in the rural areas in the upcoming decade (Rogne and Syse 2017, UNECE 2017). Centralization carries the obvious costs of longer travel time for residents of rural areas, but it may also disproportionally hit those of worst health or lower socioeconomic status. It is possible, however, that such centralization could also increase quality of some kinds of services (Goodman et al. 1997, Gooiker et al. 2011, Takahashi et al. 2021, Fischer et al. 2022), making it very involved to assess the longer-term social distribution of costs and benefits of centralizing policies.

GP is gatekeeper to specialist services in Norway, and barriers to utilization of GP may thus also affect access to other services. Specialist services are typically far more centralized than the GPs, making travel time a larger concern for utilization of such services than GPs. It also implies that GPs in rural areas may undertake more procedures than GPs who can easily refer the patient to a close-by specialist service. How this may affect the access and quality of the services is not clear, though it is well-established that volume can be important for the quality of some procedures (Luft et al. 2007, Chukmaitov et al. 2008). The question of ultimate interest is thus how travel time to the services, both GP and other, affect utilization, quality and in the end health. The trade-off between easy access (requiring i.e. short travel time) and volume of similar patients to ensure higher quality (requiring centralization) is thus difficult, with conclusions differing across types of services and political preferences.

### Strengths and limitations

The major strengths of our study included the ability to calculate the exact travel time from the resident’s home to the office of the resident’s assigned general practitioner for the Norwegian population, and to follow the residents over several years to see how starting to use the GP affected their choice of GP and residential relocation. The nation-wide registry data involves very limited or no potential for health-related attrition of participants or recall-bias, which typically influences results from studies based on self-reported data. The individual-level data allowed for adjusted and subgroup-analyses.

Still, there are several important limitations. First, we lacked exact geographic coordinates for a number of GP offices, though main findings did not change substantially when we imputed their coordinates (see Appendix). Second, it is possible that the registered home address of a resident is not representative of where he or she spend most of the time, or that travel time from work is just as relevant as travel time from home. In particular, it is well-known that students may in fact be residing in another dwelling than registered (partly due to prescriptions in the regulations), which may be the main reason for us calculating much longer travel time for students, typically in their twenties, than the rest of the population. Also, elderly or others being admitted to a long-term health care facilities typically remain registered at their home address, and, moreover, residents of such facilities often receive primary physician services in the facility instead of from their GP. Thus, results for the oldest residents, who more often live in such facilities, may be less reliable. Third, calculated travel time depends on the quality of involved algorithms using data on maps of roads and speed limits. We were able to check that the correlation between the travel times and the Pythagorean straight line, which can be calculated without involved algorithms and data on maps and speed limits, was very high (see Appendix). Fourth, a resident with a long travel distance to the GP may instead use emergency rooms, and it may thus be the travel time to the GP *relative* to the travel time to the nearest emergency room that affect utilization of GP-services and choices of residential relocation when primary physician services are needed. Since the emergency rooms in Norway are typically more centralized than GPs (Raknes et al. 2013, Legevaktutvalget 2021), this may not be an important limitation. Residents in rural areas may also have longer travel time to specialist services or private alternatives not reimbursed by the governmental health insurance (very rare in Norway), while they may use electronic/video-consultations more (not included in our measure of GP visit, but very rare in this period cf. Norwegian directorate of health 2022). In rural areas there have been problems recruiting and maintaining physicians in the GP-offices, possibly making the travel time to the office an unreliable indicator of actual distance to primary physician services (EY and Vista Analyse 2019). Fifth, analyses with higher time granulation than the year level used by us, could be undertaken to explore further whether starting to use the GP services occurred before or after the residential relocation. Also, to interpret our estimate causally, which we have been reluctant to do, we need to assume that the travel time for those who started to use the GP in the third year would have developed in the same way in the following years as it did for those who did not start to use the GP in the third year (Angrist and Pischke 2009), which is a strong assumption that may not hold. Sixth, starting to visit the GP may occur at the same time as other changes in life, possibly related to health. The motivation for relocation may thus be related to access to other amenities associated with shorter travel time to the GP, like proximity to emergency-rooms, specialist services, pharmacies, neighbors and relatives, or even other things like easily maintained condominiums instead of an older house in the countryside.

## Conclusion

Even in the sparsely populated country of Norway, travel time by car to the general practitioner (GP) was short for the vast majority of the population, suggesting that centralization of these services in urban areas are unlikely to seriously affect utilization at the population level. However, residents moved closer to the GP when they started to use the GP-services, especially patients who used to live far away. This residential relocation may reflect strong preferences for proximity to the services, and we conclude that more knowledge is needed to enable transparent balancing of costs and benefits of centralizing GP-services, at least in rural areas.

## Data Availability

The datasets that support the findings of this study contain sensitive information and are not publicly available due to privacy laws. Individual-level data for research are generally available within Norway upon application conforming with strict regulations and procedures.

### Appendix

The Population Registry data 2017 contained 5,243,697 residents, but the exact geographical location (coordinates) of their GP could only be identified for 4,067,479 of them. The dataset that could be used in the analysis (i.e. where travel time was calculated) comprised 3,976,910 residents, the difference between 4,067,479 and 3,976,910 being due to a few residents not registered with a GP and a few without sensible coordinates.

Thus, we lost more than a million residents due to lacking the coordinates of the GP. As a robustness check, we therefore imputed the location of the GP-office by giving it the median x- and y-coordinates of the x- and y-coordinates of the home of the residents assigned to the specific GP. This allowed us to calculate travel-time for 5,058,724 residents, and the observable characteristics of this extended study population was similar to those of the study population applied in the main analysis; see Appendix Table A1, second column. Travel time was, however, somewhat longer, with a median of 5.1 minutes (vs. 4.9) and a travel time at the 90^th^ percentile of 19.3 minutes (vs. 18.3); compare Appendix Figure 1 and Figure 1.

As a robustness check, we also calculated the Pythagorean straight line (in kilometers) between the coordinates of the home and the GP of each resident (using the study population of 3,976,910 residents). This straight line could be calculated without using neither the road maps nor the algorithm for travel time necessary for the analysis in minutes in the main body of the paper. The straight line and the travel time were highly correlated, with a very statistically significant (p-value <0.0000) correlation coefficient of 0.84.

**Appendix Table A1:** Summary statistics for the applied sample, as well as for the sample with imputed GP location. 2017.

|  | Sample applied i.e. with travel time to GP available | Sample with imputed GP location |
| --- | --- | --- |
| Women | 50% | 50% |
| Years of age (average) | 40.1 | 40.4 |
| Age groups |  |  |
| 0-19 | 23% | 23% |
| 20-29 | 14% | 13% |
| 30-49 | 28% | 27% |
| 50-66 | 21% | 21% |
| 67-79 | 11% | 11% |
| 80+ | 5% | 5% |
| Immigrant background | 17% | 16% |
| County of residence |  |  |
| Østfold | 6% | 6% |
| Akershus | 11% | 11% |
| Oslo | 13% | 12% |
| Hedmark | 4% | 4% |
| Oppland | 4% | 4% |
| Buskerud | 6% | 5% |
| Vestfold | 5% | 5% |
| Telemark | 4% | 3% |
| Aust-Agder | 2% | 2% |
| Vest-Agder | 3% | 4% |
| Rogaland | 9% | 9% |
| Hordaland | 10% | 10% |
| Sogn og Fjordane | 2% | 2% |
| Møre og Romsdal | 3% | 5% |
| Sør-Trøndelag | 7% | 6% |
| Nord-Trøndelag | 2% | 3% |
| Nordland | 4% | 5% |
| Troms | 3% | 3% |
| Finnmark | 1% | 1% |
| Visited GP in 2017 |  |  |
| No | 34% | 35% |
| Once | 19% | 19% |
| More than once | 47% | 46% |
| Number of residents | 3,976,910 | 5,058,724 |

**Appendix Figure A1:**
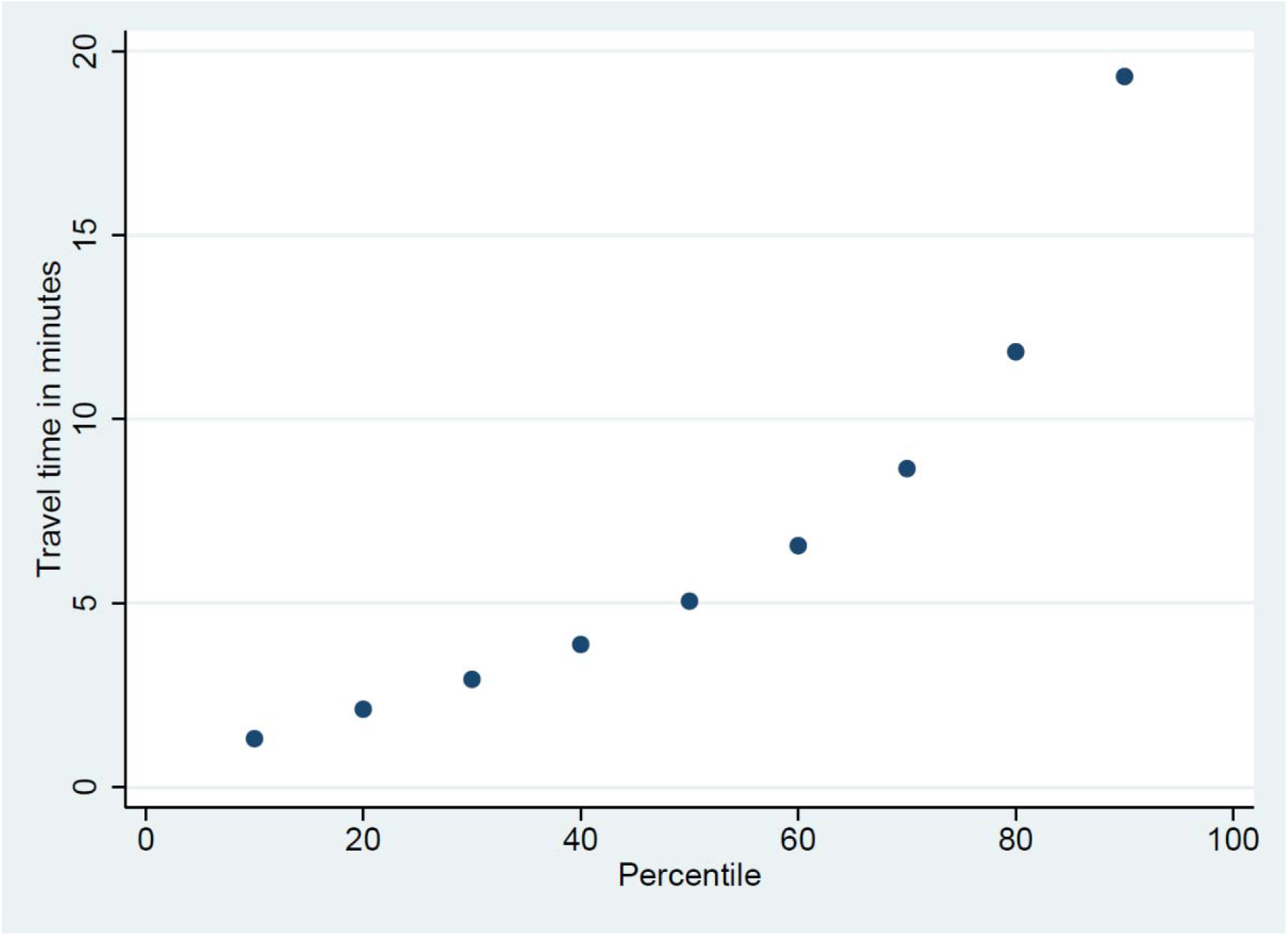
Travel time in minutes from home to the general practitioner by car for the 1^st^ to the 9^th^ decile of Norwegian residents. 2017. Based on the sample with imputed location of the GP-offices with missing coordinates. Note: The figure is the same as Figure 1, except that here the dataset also included the travel time from home to the general practitioner for the residents for whom the location of the office of the general practitioner had to be imputed.

